# The health impact of wood dust in Kijishi: Yamanaka Kijishi Study

**DOI:** 10.1101/2023.05.18.23290130

**Authors:** Yuki Nakao, Kazuhiro Okada, Yasuhiro Goto, Yosuke Matsuda, Kiyohiko Fujimoto, Yoshiyasu Okuhara

## Abstract

**Objectives:** It is well known that exposure to wood dust can cause Sinonasal carcinomas (SNC) and various respiratory diseases such as allergies. Japanese woodturners called kijishi work in an environment where wood dust is generated from lathes, and they are considered at risk for various diseases. However, there is a dearth of research on the health impacts of wood dust on kijishi. The purpose of this study is to understand the real situation of kijishi that are exposed to wood dust.

**Methods:** A questionnaire survey was conducted with 20 kijishi. We measured the amounts of wood dust two experienced kijishi were exposed to during a typical woodturning session.

**Results:** Approximately half the participants exhibited allergic symptoms such as rhinitis; however, this rate was equivalent to the result of the Japan Comprehensive Survey of Living Conditions 2019. SNC was not observed in any of the participants. The level of exposure to wood dust was low, that is, inhalable dust was 0.03 mg/m^3^, and total dust 0.22 mg/m^3^. The measured values were very low compared to the Occupational Safety and Health Administration (OSHA), National Institute for Occupational Safety and Health (NIOSH), American Conference of Governmental Industrial Hygienist (ACGIH), and Health and Safety Executive Control of Substances Hazardous to Health (HSE COSHH) standards.

**Conclusions:** We concluded that constant ventilation during woodturning reduces exposure to wood dust, thereby preventing harm to the health of kijishi to a considerable extent.

## INTRODUCTION

Kijishi are Japanese craftspeople specializing in tableware, including the wan (bowl) and the bon (tray) made by woodturning. Kijishi play important roles in maintaining traditional Japanese craftwork. They first design tableware mentally and then use a rokuro (traditional lathe) to carve each piece from a chunk of wood. The process of woodturning by using a rokuro is called *rokurobiki*. Yamanaka lacquerware originated in Kaga City, Ishikawa Prefecture, Japan. Yamanaka lacquerware is renowned for its woodturning products, has the largest population of kijishi in Japan, and produces more woodturning products than anywhere else in the country. ^1^ Yamanaka Woodturning Technique Preservation Association is one of the associations to which kijishi belong in Yamanaka lacquerware.

### Wood Dust, Preventing Exposure and Issues

Wood dust is generated during the woodturning process while using the rokuro, which may be a problem. Dust refers to solid particulates that are generated when a solid material is processed by a machine and is part of particulate matter floating in the air. In general, dust refers to such particulates with a size of up to 150 μm.^2^ The main components of wood dust are cellulose, hemicellulose, and lignin. Wood dust also contains many other low-molecular organic compounds. Studies have found that exposure to wood dust can cause not only respiratory diseases such as asthma and allergy but also sinonasal cancer sinonasal carcinomas (SNC), and it is classified as a Group 1 human carcinogen.^3,4^

To prevent apprentices from being exposed to wood dust, the training institute has installed ventilators and encourages apprentices to wear a face mask. However, little is known about kijishi who exhibit health problems from wood dust, such as respiratory diseases including asthma, allergies, and SNC. As a result, the training institute has been experiencing difficulty explaining such health problems to apprentices, and some apprentices are feeling anxious. While it is known that kijishi are exposed to wood dust when they engage in wood processing, little research regarding such exposure and related health impacts has been undertaken.

This study aimed to shed light on the health impacts of exposure to wood dust on kijishi engaging in the production of Yamanaka lacquerware products. The study was requested by the training institute, and it received support from them; the details are mentioned in the Declarations section.

## METHODS

### Participants

The study comprised a survey questionnaire and wood dust collection and measurement. Participants were members of The Yamanaka Woodturning Technique Preservation Association. Many of those who had been engaged in *rokurobiki* for a long time belong to the association. A paper-based questionnaire was sent to all members of the association individually from March 4 to 30, 2020. No compensation was given for completing the questionnaire.

### Questionnaire Survey

A questionnaire survey was administered to 20 kijishi who were members of the association. The questionnaire asked participants about items shown in Table 1.

**Table 1.** Questionnaire items

|  |  |
| --- | --- |
| Participants' background information | Gender, age, number of years of experience as kijishi, number of hours of work per day, history of occupation other than kijishi |
| Presence or absence of disease | Presence or absence of allergic diseases such as rhinitis and cough (symptoms including shortness of breath, nasal congestion, rhinorrhea, and headache); presence or absence of sinonasal cancer/ diabetes/hypertension/dyslipidemia/lower back pain/hearing impairment, as well as related past history, medication history, and family history |
| Lifestyle habits of participants | Presence or absence of alcohol consumption, smoking, and exercise habit |
| Information on the work environment | Use of a face mask during woodturning, availability and use of ventilation equipment during woodturning |

### Dust Measurement

In this study, levels of exposure to wood dust were measured with support from two kijishi (with 41 and 39 years of experience) who were both members of the association.

### Wood and Conditions for Rokurobiki

Sugi (*Cryptomeria japonica*) is commonly used for building material in Japan; however, it is not suitable for woodturning because the growth rings between its latewood (winter) and earlywood (summer) bands are made of soft fibers that break during woodturning.^5^ Hence, kijishi rarely choose sugi for woodturning. Instead, they tend to use keyaki (*Zelkova serrata*) as it can withstand the lathe and has a beautiful texture. Therefore, this study used chunks of dried keyaki (after rough turning) with a water content of 10.1% (measured by comparing samples taken before and after drying). Measurements were taken in a typical kijishi work environment (Figure 1-a). The rokuro was operated at a maximum of 1000 rpm.

**Figure 1.** Work scene, etc. 1-a: Working on Rokurobiki and dust measurement in the workshop 1-b: Schematic diagram of the workshop 1-c: Work with the door slightly open. 1-d: Generation of wood dust through the use of coated abrasives

### Conditions for Ventilation

Kijishi work as sole traders and have their own workshops. They have ventilation equipment installed in their workshops, such as a dust collector and a ventilation fan. Most kijishi adjust the settings of ventilation equipment depending on the amount of dust being generated. In this study, wood dust was measured while ventilation equipment was being operated because 1) almost all kijishi who participated in the questionnaire survey were using ventilation equipment; and 2) kijishi would have been at risk of being exposed to wood dust if the ventilation equipment had not been operated.

Additionally, it was found that most kijishi secure an air inlet by opening windows located upstream of the ventilation equipment so that the air flows effectively. As such, the ventilation equipment was in operation, and the sliding door located upstream of the equipment was left ajar during dust measurement. We confirmed that the wind velocity was 0.1 m/s or lower at the dust collection point before measurement (Figure 1-b, 1-c).

### Wood Dust Collection Methods

Dust particle sizes and measurement methods are key in measuring wood dust.

In this study, particle-size distribution, inhalable dust, and total dust were measured.

An Andersen-type air sampler (Sibata Scientific Technology Ltd., AN-200) was used to measure particle-size distribution. The Andersen-type air sampler collects air samples that include dust at an inhalation flow rate of 28.3 L/min, and (according to the package insert) can classify collected samples into the following nine levels by aerodynamic particle size: ≤0.43 μm, 0.43–0.65 μm, 0.65–1.1 μm, 1.1–2.1 μm, 2.1–3.3 μm, 3.3–4.7 μm, 4.7–7.0 μm, 7.0–11.0 μm, and ≥11.0 μm. Thereafter, the masses of the dust samples (mg) were measured for each particle size to obtain mass-based particle sizes.

The AN-200 was placed 65 mm away from the center of the bowl toward the kijishi and at a height of 590 mm above the workbench. According to the questionnaire, kijishi spend an average of approximately 8 hours per day on woodturning. Taking into account that the room is sufficiently ventilated during a lunch break, we considered that kijishi are consecutively exposed to wood dust for a mean duration of 4 hours per day. Hence, wood dust for measurement was collected for 4 hours in this study (Figure 1-d). The participating kijishi carried out tasks in 2-hour shifts. Eleven woodturning pieces were completed over the course of 4 hours.

An inertial impaction-type air sampler (Sibata Scientific Technology Ltd., NW-354) was used for measuring inhalable dust and total dust. Inertial impaction-type air samplers enable size separation and collection of particles in the air by inertial impaction.^6^ Total dust refers to dust collected at a velocity of 0.5–0.8 m/s at the inlet of the dust collecting devices.^7^ Inhalable dust is defined as dust that penetrates a dust sampler with the following size separation properties: a collection efficiency of 50% for dust with a size of up to 4.0 μm, and 0% for dust with a size of 10 μm or greater^7^ (Table 2).

**Table 2.** Types and definitions of dust

|  |  |
| --- | --- |
| Dust | Solid particulates that are generated when a solid material is processed by machine. These solid particulates are part of particulate matter floating in the air. |
| Inhalable dust | Dust that penetrates a sampler with the following size separation properties: a collection efficiency of 50% for dust with a size of up to 4.0 µm; and a collection efficiency of 0% for dust with a size of 10 µm or greater |
| Total dust | Dust that is collected at a velocity of 50–80 cm/sec at the inlet of the collecting device |

The NW-354 was placed on the opposite side of the kijishi, 100 mm away from the center of the bowl, at a height of 460 mm above the workbench. Wood dust for measurement was also collected for a duration of 4 hours.

Also, background data were measured inside the room while woodturning was not being performed. Samples for background data were collected under the same conditions as those for measuring dust, that is, for 4 hours at a wind velocity of up to 0.1 m/s at the dust collection point.

## RESULTS

### Questionnaire Results (Table 3)

We collected responses from all 20 kijishi surveyed. The mean age of participants was 59.9 years old (±13.6). Only one woman participated. Three participants had previously worked in occupations other than kijishi, namely as a gas station worker, furniture designer, and car mechanic, none of which involve exposure to wood dust. The history of asbestos inhalation and other related information for the three participants could not be confirmed.

**Table 3.**
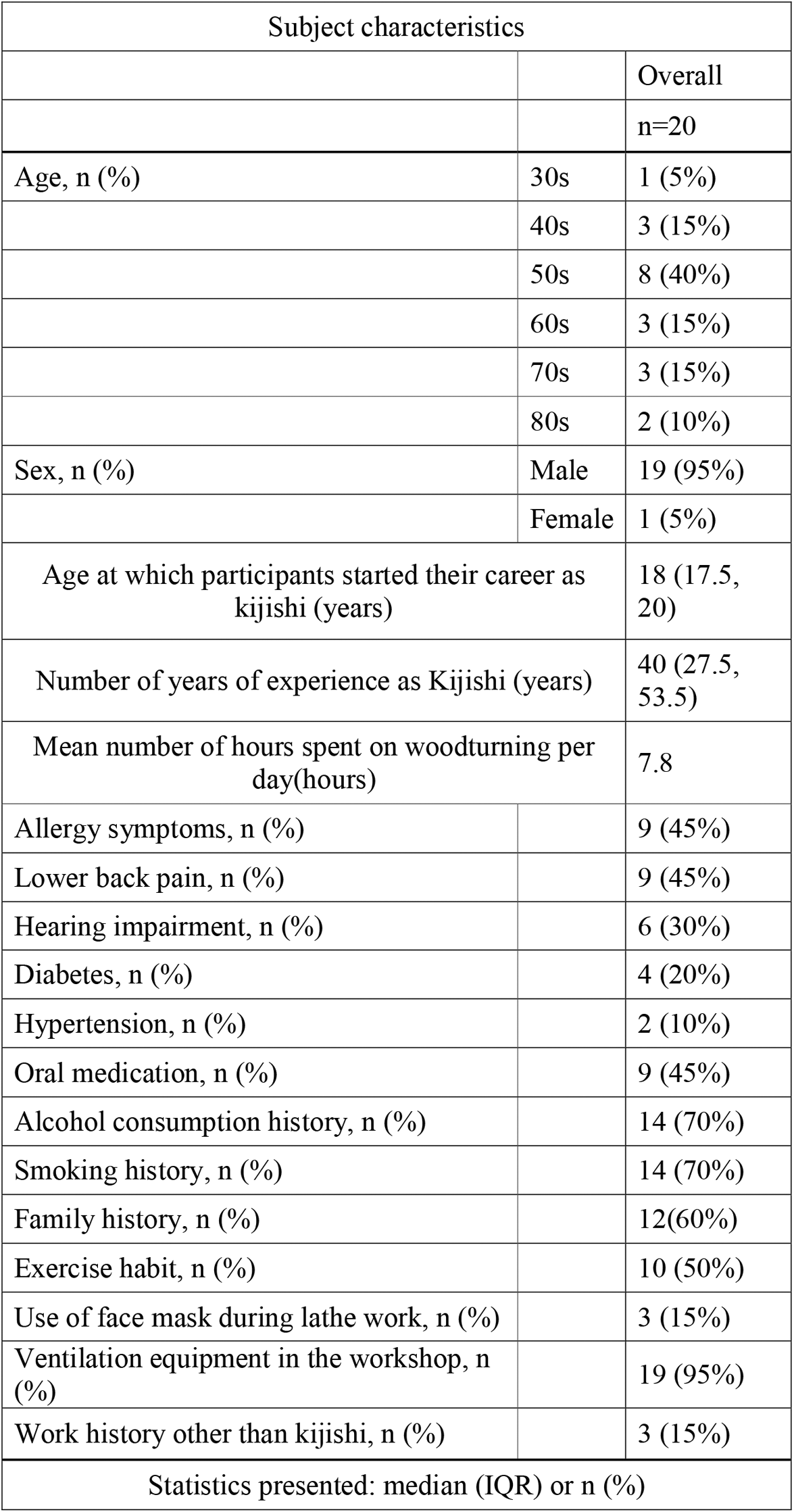
Questionnaire results: Subject characteristics

Nine participants indicated the presence of diseases, including lower back pain and allergy symptoms, such as rhinitis and coughing. Four participants had diabetes, and two had hypertension. None of the participants had SNC. Nine participants were taking medications, including analgesics, agents for improving uric acid levels and those for improving liver function, and antidiabetic and antihypertensive agents. Regarding family history, four participants had a family history of lung cancer, two of gastrointestinal cancer, and six of other cancers (details unknown).

As for lifestyle habits, 14 were consuming alcohol and smoking cigarettes. Ten participants were regularly engaging in exercise, such as walking. As for the work environment, only three participants reported using a face mask during woodturning. However, all but one of the participants had a room ventilator installed inside the workshop and were using it during woodturning. Data were considered to have been normally distributed according to the histogram (Figure S1), and median (IQR) or n (%) were used in indicating age (Table 3). As participants included only one woman, a histogram created by gender would have revealed her age. Therefore, the histogram indicates only the age distribution of all participants and the number of participants in each age group.

### Wood Dust Measurement Results

Background data were collected at a room temperature of 25.7°C and humidity of 67.8%. Data for dust measurement were collected at a room temperature of 26.9°C and humidity of 62.8%. The particle size distribution of dust could not be calculated as only a limited amount of dust could be collected. Inhalable dust in the background and during woodturning was 0.04 mg/m^3^ and 0.03 mg/m^3^, respectively. Total dust in the background and during woodturning was 0.04 mg/m^3^ and 0.22mg/m^3^, respectively.

## DISCUSSION

As for inhalable dust, the measured value of 0.03 mg/m^3^ during woodturning was almost the same as the background value of 0.04 mg/m^3^, and the smaller value during woodturning was probably due to measurement error. This may show that the dust concentration during woodturning is very small within the error margin. This indicates that the Kijishi were adequately ventilated during woodturning and that their exposure to inhalable dust was very low.

It is well known that exposure to wood dust can cause a host of illnesses, including reduced respiratory function, lung disease, asthma, and even SNC. The Japanese government has created a list of allergens that can cause allergic respiratory disease (e.g., allergic rhinitis, bronchial asthma, pharyngitis), including western red cedar (*Thuja plicata*), nezuko (*Thuja standishii*), lauan, ryobu (*Clethra barbinervis*), mulberry, hoonoki (*Magnolia obovate*), and Shirakaba (Betula *platyphylla*). Allergic respiratory disease caused by exposure to wood dust of these types of wood is treated as an occupational disease in the country.^8^ Although keyaki (*Zelkova serrata*) is not on the list, it can also cause allergic respiratory disease.

In this study, approximately half of the participants indicated that they had allergic symptoms such as rhinitis. All of those who responded to the survey as such were male. This percentage was almost the same as the results of the latest national epidemiological survey in Japan (the percentage of men with allergy symptoms: 49.7% in 2019).^9^ Since this survey was conducted in March, which generally corresponds to the time when allergic rhinitis symptoms caused by Sugi (*Cryptomeria japonica*) and Hinoki (*Chamaecyparis obtusa*) are most likely to appear, the percentage of complaints of allergic symptoms obtained in this survey was also considered to be influenced by those pollens.

It is known that there is a positive association between smoking and allergy onset.^10^ Fourteen of the twenty participants (four were smokers and ten had smoked in the past) had a history of smoking, which may have also influenced the proportion of participants reporting allergic symptoms.

As noted previously, most particles with a particle size of 10 μm or greater remain in the nasal cavity and throat and are swallowed or spat in the form of phlegm.^11^ We found that most dust particles collected during woodturning were of this size (i.e., larger than inhalable dust). Such particles remain in the nasal cavity leading to allergic symptoms such as rhinitis. Wood Dust Exposure Limits are specified by the respective agencies as follows: ^8^

a. Occupational Safety and Health Administration (OSHA): total dust =15 mg/m^3^, respirable fraction = 5 mg/m^3^ or less;
b. National Institute for Occupational Safety and Health (NIOSH): total dust = 1 mg/m^3^ or less;
c. American Group of Governmental Industrial Hygienist (AGGIH): Inhalable particulate matter = 0.5 mg/m^3^ for western red cedar and inhalable particulate matter = 1 mg/m^3^ or less for other than western red cedar;
d. Health and Safety Executive Control of Substances Hazardous to Health (HSE COSHH) essentials (2020) stipulates: inhalable fraction = 3 mg/m^3^ or less.

We in the present study found that the level of exposure to wood dust was low, that is, inhalable dust was 0.03 mg/m^3^, and total dust 0.22 mg/m^3^. The measured values were very low compared to OSHA, NIOSH, ACGIH, and HSE COSHH standards.

Some participants indicated that they experienced coughs and other symptoms when they inhaled large amounts of wood dust while making numerous large wooden items (e.g., Japanese round tea box) in the 1970s. In those times, ventilation equipment was yet to be installed; therefore, large amounts of wood dust were suspended in the air. This finding is likely because constant ventilation during woodwork reduces exposure to wood dust.

As noted, none of the participants indicated the presence of SNC, which is a rare disease with morbidity of 1–2 in 100,000. In recent years, the number of patients with SNC has begun declining even further in Japan.^12^ Data disclosed by the Cancer Information Center of the National Cancer Center do not include information on malignant neoplasms of the nasal cavity or the paranasal sinuses. The latest morbidity rate of these diseases is unknown. Researchers considered that no participants in this study had SNC because it is an extremely rare disease. Hardwoods are considered to have a high risk of SNC.^13^ Although there are no reports on the carcinogenicity of keyaki (*Zelkova serrata*), it is considered necessary to be careful about the occurrence of SNC considering that keyaki (*Zelkova serrata*) is a hardwood. Surveying a larger sample of kijishi could reveal cases of SNC.

### Limitations

This study relied on a questionnaire survey with a small sample size; therefore, the carcinogenicity of wood dust cannot be discussed on the basis of the present findings. Kijishi with advanced woodturning skills only require small amounts of coated abrasives during rokurobiki. Therefore, experienced kijishi are unlikely to be exposed to large amounts of wood dust.

Dust should be measured at the position of the nose of the kijishi. However, the measurement device could not be placed at the nose position as doing so would have interfered with tasks.

Further, it should be noted that this study has a selection bias because some kijishi do not belong to the Yamanaka Woodturning Technique Preservation Association.

## CONCLUSION

The real status of kijishi exposed to wood dust in the Yamanaka Woodturning Technique Preservation Association was revealed. Since the amount of wood dust exposure in the kijishi-workshops was very small due to the use of ventilation equipment, the potential health effects of wood dust were considered to be small.

Due to the small sample size of this survey, we believe that this study can be used as a starting point for a more detailed study through a large-scale survey of kijishi involved in lacquerware production in Japan.

## Data Availability

All data produced in the present work are contained in the manuscript

## DATA SHARING AND DATA ACCESSIBILITY

The datasets used and/or analyzed during the current study are available from the corresponding author on reasonable request.

## ACKNOWLEDGMENTS

We would like to thank all the kijishi of the Yamanaka Woodturning Technique Preservation Association who participated in the survey.

We thank Shoichi Mukaide, Hirohiko Kawakita and Ryozo Kawagita for their guidance and work.

Moreover, the authors extend their sincere gratitude to all of those who spent their valuable time participating in this study. This study received financial support from the Public Interest Incorporated Foundation Japan Foundation for Promoting Welfare of Small and Medium-sized Enterprises (FY2020 Nihon Full Happ Research Grant).

## DISCLOSURE

### Approval of the Research Protocol

The study was approved by the ethics review committee of the Kaga Medical Center as well as the Yamanaka Woodturning Technique Preservation Association (approval number R1-8).

### Informed Consent

Written consent was obtained from all participants.

### Registry and the Registration No. of the study/trial

N/A.

### Animal Studies

N/A.

## Supporting Information

**Figure S1**. A histogram of participants age groups (years)

