## Supplementary figures and images for "The health impact of wood dust in Kijishi: Yamanaka Kijishi Study"

### Supporting Information

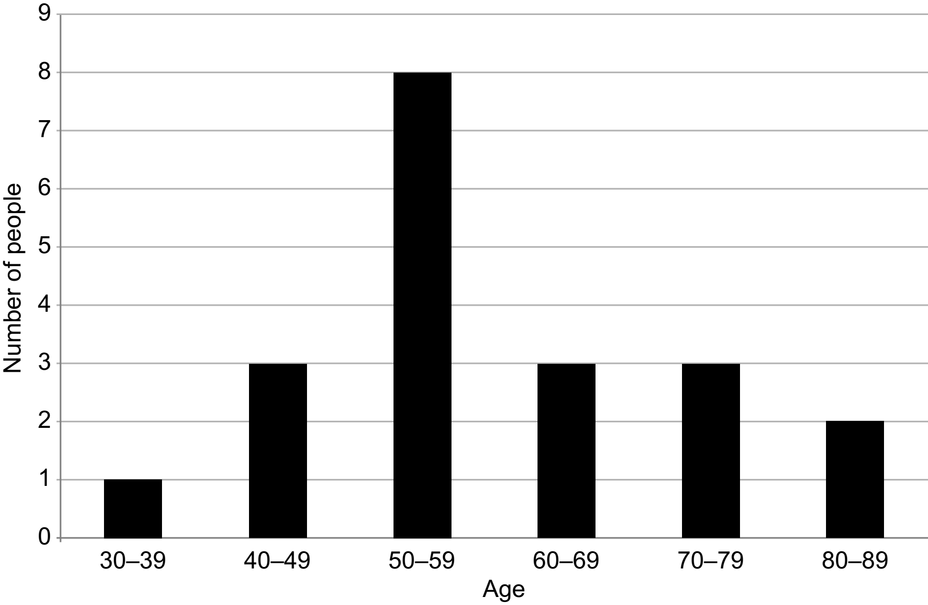
